# IMPACT OF CONDITIONING INTENSITY AND GENOMICS ON RELAPSE AFTER ALLOGENEIC TRANSPLANTATION FOR PATIENTS WITH MYELODYSPLASTIC SYNDROME

**DOI:** 10.1101/2020.08.25.20138461

**Authors:** Laura W. Dillon, Gege Gui, Brent R. Logan, Mingwei Fei, Jack Ghannam, Yuesheng Li, Abel Licon, Edwin P. Alyea, Asad Bashey, Steven M. Devine, Hugo F. Fernandez, Sergio Giralt, Mehdi Hamadani, Alan Howard, Richard T. Maziarz, David L. Porter, Erica D. Warlick, Marcelo C. Pasquini, Bart L. Scott, Mitchell E. Horwitz, H. Joachim Deeg, Christopher S. Hourigan

## Abstract

Myelodysplastic Syndrome (MDS) patients are at risk of relapse after allogeneic hematopoietic cell transplantation (alloHCT). The utility of ultra-deep genomic testing to predict, and the impact of conditioning intensity to prevent, MDS relapse are unknown. Targeted error-corrected DNA sequencing was performed on pre-conditioning blood samples from the Blood and Marrow Transplant Clinical Trials Network (BMT CTN) 0901 phase III randomized clinical trial which compared outcomes by alloHCT conditioning intensity in adult patients with less than 5% marrow myeloblasts and no leukemic myeloblasts in blood on morphological analysis at the time of pre-transplant assessment. Using a previously described set of 10 gene regions, 42% of patients had mutations detectable prior to randomization to reduced intensity or myeloablative conditioning. Testing positive was associated with increased rates of relapse and decreased overall survival. In those testing positive, relapse rates were higher and relapse-free survival was lower in reduced intensity versus myeloablative conditioning arms. Testing additional genes, including those associated with MDS, did not improve prognostication. This study provides evidence that post-transplant relapse rates in MDS patients are highest in those with pre-transplant genomic evidence of high-risk disease. In those testing positive, randomization to myeloablative conditioning lowered but did not eliminate relapse risk.

## Introduction

Myelodysplastic syndrome (MDS), one of the most common hematologic disorders, is a collection of clinically and genetically heterogenous diseases. Allogeneic hematopoietic cell transplantation (alloHCT) is currently the only curative treatment for MDS but usage is limited, in part, by the risk of transplant-related mortality (TRM).^1^ Reduced intensity conditioning (RIC) regimens have helped to decrease toxicity although multiple retrospective studies found that the decreased TRM is counterbalanced by increased risk of relapse compared with more intense myeloablative conditioning (MAC) regimens.^2,3^ Two randomized phase III trials comparing outcomes following RIC and MAC conditioning regimens in MDS patients have yet to provide a definitive answer regarding which regimen should be used when a patient is eligible for either approach.^4-6^

Aside from conditioning regimen, a variety of other factors, including the presence and type of genetic mutations before and after transplant, have been found to influence clinical outcomes.^7-9^ We recently demonstrated that acute myeloid leukemia (AML) patients with detectable mutations prior to alloHCT had decreased relapse and improved overall survival when randomized to MAC versus RIC.^10^ To determine if a similar benefit from increased conditioning intensity is seen for MDS patients we performed ultra-deep error-corrected DNA sequencing on blood samples collected immediately prior to random assignment to either MAC or RIC for alloHCT.

## Methods

### Clinical Cohort

The Blood and Marrow Transplant Clinical Trials Network (BMT CTN) 0901 (NCT01339910) study was a phase III randomized clinical trial comparing outcomes by conditioning intensity in adult patients with myeloid malignancy undergoing alloHCT with less than 5% marrow myeloblasts and no leukemic myeloblasts in blood on morphological analysis at the time of pre-transplant assessment.^6^ Frozen whole blood collected prior to the conditioning regimen was available from 48 of the 54 MDS patients. Samples came from patients receiving either MAC (n=25) or RIC (n=23); these groups were well matched for baseline characteristics (**Table 1**). Extended follow-up of patients enrolled on this protocol was extracted from the Center for International Blood and Marrow Transplant Research (CIBMTR) research database. Median follow-up in survivors was in excess of 53 months. Patient characteristics and clinical outcomes were aligned with those previously reported (**supplemental Figure 1**). Patients provided written informed consent to participate in both the BMT CTN 0901 trial and the CIBMTR research database. This study was approved by the BMT CTN and CIBMTR and conducted with approval of the National Marrow Donor Program institutional review board.

**Table 1.** Patient Clinical Characteristics.

| **Characteristic** | **MAC** | **RIC** | **Total** |
| --- | --- | --- | --- |
| Number of patients | 25 | 23 | 48 |
| **Age** |  |  |  |
| Median (range) | 58.1 (26-65.9) | 55.2 (24.1-63.6) |  |
| ≤50 | 3 (12%) | 10 (43.5%) | 13 |
| >50 | 22 (88%) | 13 (56.5%) | 35 |
| **Gender** |  |  |  |
| Female | 10 (40%) | 9 (39.1%) | 19 |
| Male | 15 (60%) | 14 (60.9%) | 29 |
| **HCT-CI** |  |  |  |
| 0 | 8 (32%) | 4 (17.4%) | 12 |
| 1-2 | 8 (32%) | 13 (56.5%) | 21 |
| >2 | 9 (36%) | 6 (26.1%) | 15 |
| **Disease** |  |  |  |
| Refractory Anemia | 3 (12%) | 1 (4.3%) | 4 |
| Refractory Anemia with Ringed Sideroblasts | 3 (12%) | 2 (8.7%) | 5 |
| Refractory Cytopenia with Multilineage Dysplasia | 4 (16%) | 8 (34.8%) | 12 |
| Refractory Cytopenia with Multilineage Dysplasia and Ringed Sideroblasts | 0 (0%) | 1 (4.3%) | 1 |
| Refractory Anemia with Excess Blasts - 1 (5-9% blasts) | 4 (16%) | 6 (26.1%) | 10 |
| Refractory Anemia with Excess Blasts - 2 (10-19% blasts) | 5 (20%) | 4 (17.4%) | 9 |
| MDS, Unclassified | 4 (16%) | 1 (4.3%) | 5 |
| MDS Associated with Isolated Del(5q) | 2 (8%) | 0 (0%) | 2 |
| **Disease risk** |  |  |  |
| Standard | 19 (76%) | 16 (69.6%) | 35 |
| High | 6 (24%) | 7 (30.4%) | 13 |
| **Disease duration (months)** |  |  |  |
| Median (range) | 7.7(2.7-70.3) | 11.4(1.5-129.2) |  |
| 0-2.9 | 2 (8%) | 1 (4.3%) | 3 |
| 3-5.9 | 6 (24%) | 6 (26.1%) | 12 |
| 6-11.9 | 10 (40%) | 5 (21.7%) | 15 |
| ≥12 | 7 (28%) | 11 (47.8%) | 18 |
| **Cytogenetics** |  |  |  |
| Favorable | 10 (40%) | 8 (34.8%) | 18 |
| Intermediate | 4 (16%) | 8 (34.8%) | 12 |
| Poor | 11 (44%) | 7 (30.4%) | 18 |
| **Donor type** |  |  |  |
| Related | 10 (40%) | 8 (34.8%) | 18 |
| Unrelated | 15 (60%) | 15 (65.2%) | 30 |
| **Donor match** |  |  |  |
| Matched | 25 (100%) | 23 (100%) | 48 |
| Mismatched | 0 (0%) | 0 (0%) | 0 |
| **Graft type** |  |  |  |
| PB | 23 (92%) | 21 (91.3%) | 44 |
| BM | 2 (8%) | 2 (8.7%) | 4 |
| **Conditioning regimen** |  |  |  |
| Flu/Bu4 | 22 (88%) | NA | 22 |
| Bu/Cy | 2 (8%) | NA | 2 |
| Cy/TBI | 1 (4%) | NA | 1 |
| Flu/Mel | NA | 6 (26.1%) | 6 |
| Flu/Bu2 | NA | 17 (73.9%) | 17 |
| **ATG** |  |  |  |
| ATG | 3 (12%) | 4 (17.4%) | 7 |
| No ATG | 22 (88%) | 19 (82.6%) | 41 |

Abbreviations: ATG, antithymocyte globulin; BM, bone marrow; Bu, busulfan; Cy, cyclophosphamide; Flu, fludarabine; Hct-CI, Hematopoietic Cell Transplant-Cormorbidity Index; ITD, internal tandem duplication; MAC, myeloablative conditioning; Mel, melphalan; MDS, myelodysplastic syndrome; NA, not applicable; PB, peripheral blood; RIC, reduced-intensity conditioning; TBI, total body irradiation.

### Sequencing

DNA sequencing using a custom anchored multiplex PCR-based panel (ArcherDx, Boulder, CO) designed to incorporate molecular barcode/unique molecular identifiers (UMIs) and cover regions of 29 genes commonly mutated in myeloid malignancies (**supplemental Table 1**) was performed on 200ng of genomic DNA isolated from each preconditioning blood sample. Library preparation and paired-end 150-bp sequencing was performed using unique dual-sample indices on a Hiseq 2500 (rapid run mode; Illumina, San Diego, CA) as previously described.^10^ An average of 43 million paired-end reads were acquired per sample (**supplemental Table 2**).

### Bioinformatic and Statistical Analyses

Sequence analysis, variant calling, and statistical analyses were performed as reported previously^10^ and are described in the supplement.

## Results

### Presence of mutations in 10 genes predicts clinical outcome

Previously we demonstrated that detection of mutations within 10 gene regions *(FLT3, IDH1, IDH2, JAK2, KIT, NPM1, NRAS, RUNX1, SF3B1*, and *TP53)* prior to alloHCT in AML patients in remission was associated with higher relapse and lower survival in those randomized to RIC.^10^ Evaluating the same gene regions in this MDS cohort, a total of 54 mutations with a median variant allele frequency (VAF) of 0.7% were detected in the blood of 42% (n = 20) of patients prior to conditioning treatment (**supplemental Figure 2 and supplemental Table 3**). In samples with mutations detected, the median number of variants was 2 (range 1-11).

The presence of a mutation in the 10 gene panel (NGS_g10_) in the blood of MDS patients prior to conditioning was found to be prognostic. NGS_g10_ positive patients experienced significantly higher rates of relapse (3-year relapse, 40% vs 11%; *P* = 0.02) and decreased relapse-free survival (3-year RFS, 34% vs 71%, *P* = 0.006) and overall survival (3-year OS, 55% vs 79%, *P* = 0.05) compared to NGS_g10_ negative patients (**Figure 1, Table 2**). NGS_g10_ mutational status served as a strong predictor of relapse in this cohort of MDS patients, predicting 73% of relapses at 24 months (100% for those receiving MAC), with a specificity of 78% (100% for those receiving RIC) (**supplemental Table 4**).

**Figure 1.**
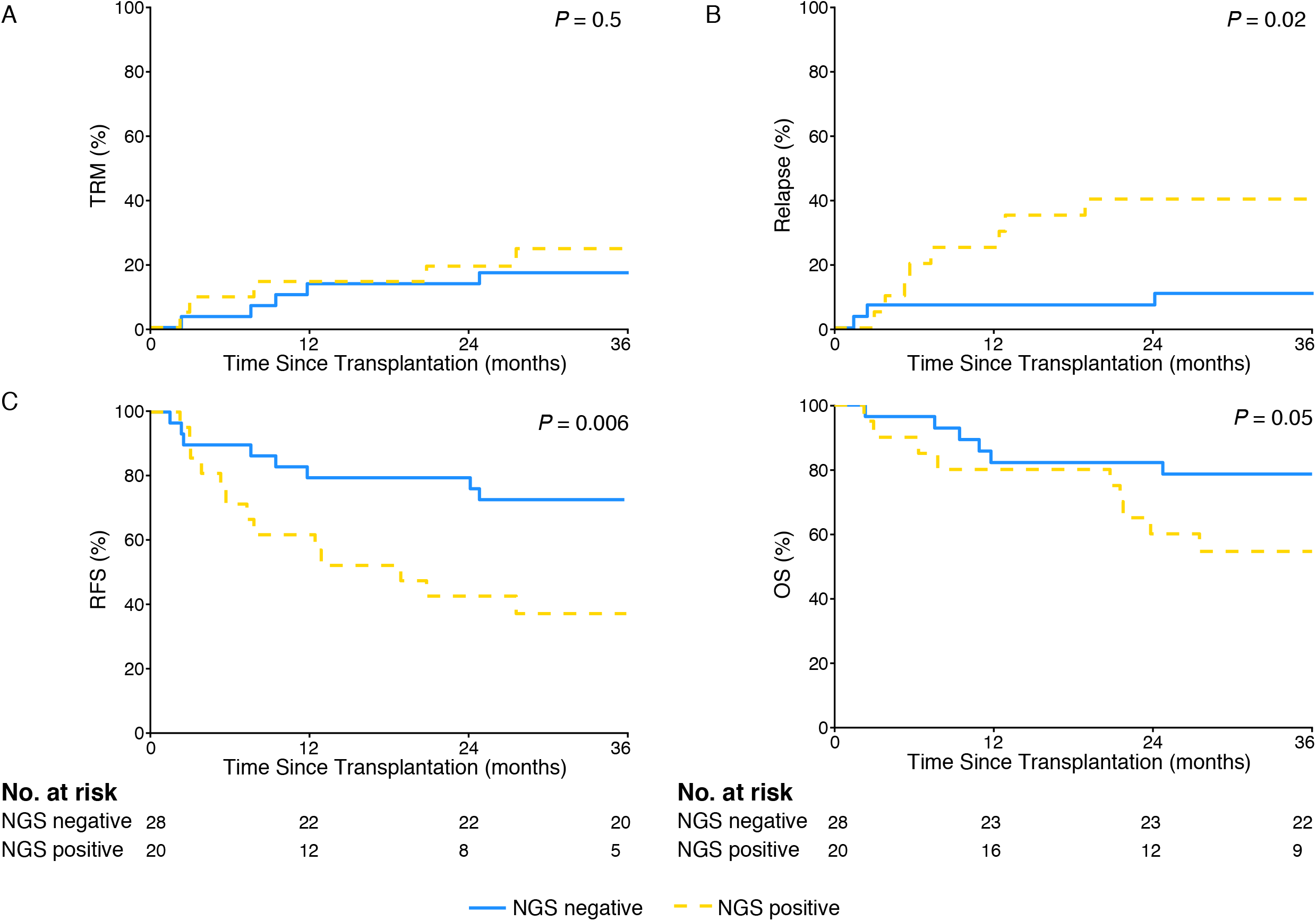
Impact of 10-gene mutational status on MDS patient clinical outcomes. (A) No difference in transplant-related mortality was observed between MDS patients based on 10-gene mutational status (P = 0.5). (B) Rates of relapse were significantly higher in 10-gene next-generation sequencing (NGS) positive versus NGS negative patients (3-year relapse 40% vs. 11%, *P* = 0.02). (C) 10-gene NGS positive patients had significantly decreased relapse-free survival (3-year RFS, 34% vs 71%, *P* = 0.006) and overall survival (3-year OS, 55% vs 79%, *P* = 0.05) compared to NGS negative patients.

**Table 2.** Outcomes analysis by 10-gene next-generation sequencing status and treatment regimen.

|  | NGS pos<br>n = 20 | NGS neg<br>n = 28 |  | MAC<br>NGS pos<br>n = 12 | MAC<br>NGS neg<br>n = 13 | RIC<br>NGS pos<br>n = 8 | RIC<br>NGS neg<br>n = 15 |  |
| --- | --- | --- | --- | --- | --- | --- | --- | --- |
| Outcome | Freq<br>(95% CI) | Freq<br>(95% CI) | *P* | Freq<br>(95% CI) | Freq<br>(95% CI) | Freq<br>(95% CI) | Freq<br>(95% CI) | *P* |
| **Treatment-related mortality** |  |  | **0.466** |  |  |  |  | **0.692** |
| 3 months | 10(0-24) | 4(0-11) |  | 8(0-25) | 8(0-23) | 13(0-37) | 0(0-0) |  |
| 6 months | 10(0-24) | 4(0-11) |  | 8(0-25) | 8(0-23) | 13(0-37) | 0(0-0) |  |
| 1-year | 15(0-31) | 14(1-28) |  | 17(0-39) | 23(0-47) | 13(0-37) | 7(0-20) |  |
| 2-year | 20(2-38) | 14(1-28) |  | 25(0-51) | 23(0-47) | 13(0-37) | 7(0-20) |  |
| 3-year | 26(5-46) | 18(3-32) |  | 35(5-64) | 23(0-47) | 13(0-37) | 13(0-31) |  |
| **Relapse** |  |  | **0.022** |  |  |  |  | **<0.001** |
| 3 months | 0(0-0) | 7(0-17) |  | 0(0-0) | 0(0-0) | 0(0-0) | 13(0-31) |  |
| 6 months | 20(2-38) | 7(0-17) |  | 0(0-0) | 0(0-0) | 50(11-89) | 13(0-31) |  |
| 1-year | 25(5-45) | 7(0-17) |  | 0(0-0) | 0(0-0) | 63(24-100) | 13(0-31) |  |
| 2-year | 40(18-62) | 7(0-17) |  | 17(0-39) | 0(0-0) | 75(38-100) | 13(0-31) |  |
| 3-year | 40(18-62) | 11(0-22) |  | 17(0-39) | 0(0-0) | 75(38-100) | 20(0-41) |  |
| **Relapse-free survival** |  |  | **0.006** |  |  |  |  | **<0.001** |
| 3 months | 90(66-97) | 89(70-96) |  | 92(54-99) | 92(57-99) | 88(39-98) | 87(56-97) |  |
| 6 months | 70(45-85) | 89(70-96) |  | 92(54-99) | 92(57-99) | 38(9-67) | 87(56-97) |  |
| 1-year | 60(36-78) | 79(58-90) |  | 83(48-96) | 77(44-92) | 25(4-56) | 80(50-93) |  |
| 2-year | 40(19-60) | 79(58-90) |  | 58(27-80) | 77(44-92) | 13(1-42) | 80(50-93) |  |
| 3-year | 34(15-55) | 71(51-85) |  | 49(19-73) | 77(44-92) | 13(1-42) | 67(38-85) |  |
| **Overall survival** |  |  | **0.045** |  |  |  |  | **0.120** |
| 3 months | 90(66-97) | 96(77-100) |  | 92(54-99) | 92(57-99) | 88(39-98) | 100 |  |
| 6 months | 90(66-97) | 96(77-100) |  | 92(54-99) | 92(57-99) | 88(39-98) | 100 |  |
| 1-year | 80(55-92) | 82(62-92) |  | 83(48-96) | 77(44-92) | 75(32-93) | 87(56-97) |  |
| 2-year | 60(36-78) | 82(62-92) |  | 67(34-86) | 77(44-92) | 50(15-78) | 87(56-97) |  |
| 3-year | 55(31-73) | 79(58-90) |  | 57(25-80) | 77(44-92) | 50(15-78) | 80(50-93) |  |

Abbreviations: NGS, next-generation sequencing; pos, positive; neg, negative; MAC, myeloablative conditioning; RIC, reduced intensity conditioning; Freq, frequency; CI, confidence interval.
Overall *P* values (in bold): Gray's test for treatment-related mortality and relapse; log-rank test for relapse-free survival and overall survival.

In contrast to NGS_g10_ mutational status, no significant difference in rates of relapse or overall survival was observed when stratifying patients by disease classification, disease risk group, or cytogenetic prognostic group (**supplemental Figure 3**). Rates of relapse trended higher in patients with poor cytogenetics or categorized as high risk. Inclusion of patients with poor cytogenetics prior to transplant with NGS_g10_ mutational status improved prediction of relapse at 24 months from 73% to 91% (**supplemental Table 4**) and was associated with significantly higher rates of relapse compared to NGS_g10_ negative patients (**supplemental Figure 4**).

### Presence of mutations in 10 genes predicts relapse by conditioning intensity

Next, the impact of conditioning intensity and NGS_g10_ mutational status was examined. Mutations were detected in the pre-conditioning blood samples of 48% of MAC and 35% of RIC patients. Patients testing positive experienced higher rates of relapse (3-year relapse, 75% RIC vs 17% MAC; *P* = 0.003) and lower relapse-free survival (3-year RFS, 13% RIC vs. 49% MAC; *P* = 0.003) when randomized to RIC rather than MAC (**Figure 2AB, Table 2**). Neither of the two relapses observed after MAC occurred in the first 12 months after transplant, the median time to relapse in those randomized to RIC was 4 months (range 2-24 months). No significant difference was observed in rates of TRM or overall survival when stratifying patients by both NGS_g10_ status and conditioning intensity (**Figure 2C, Table 2**).

**Figure 2.**
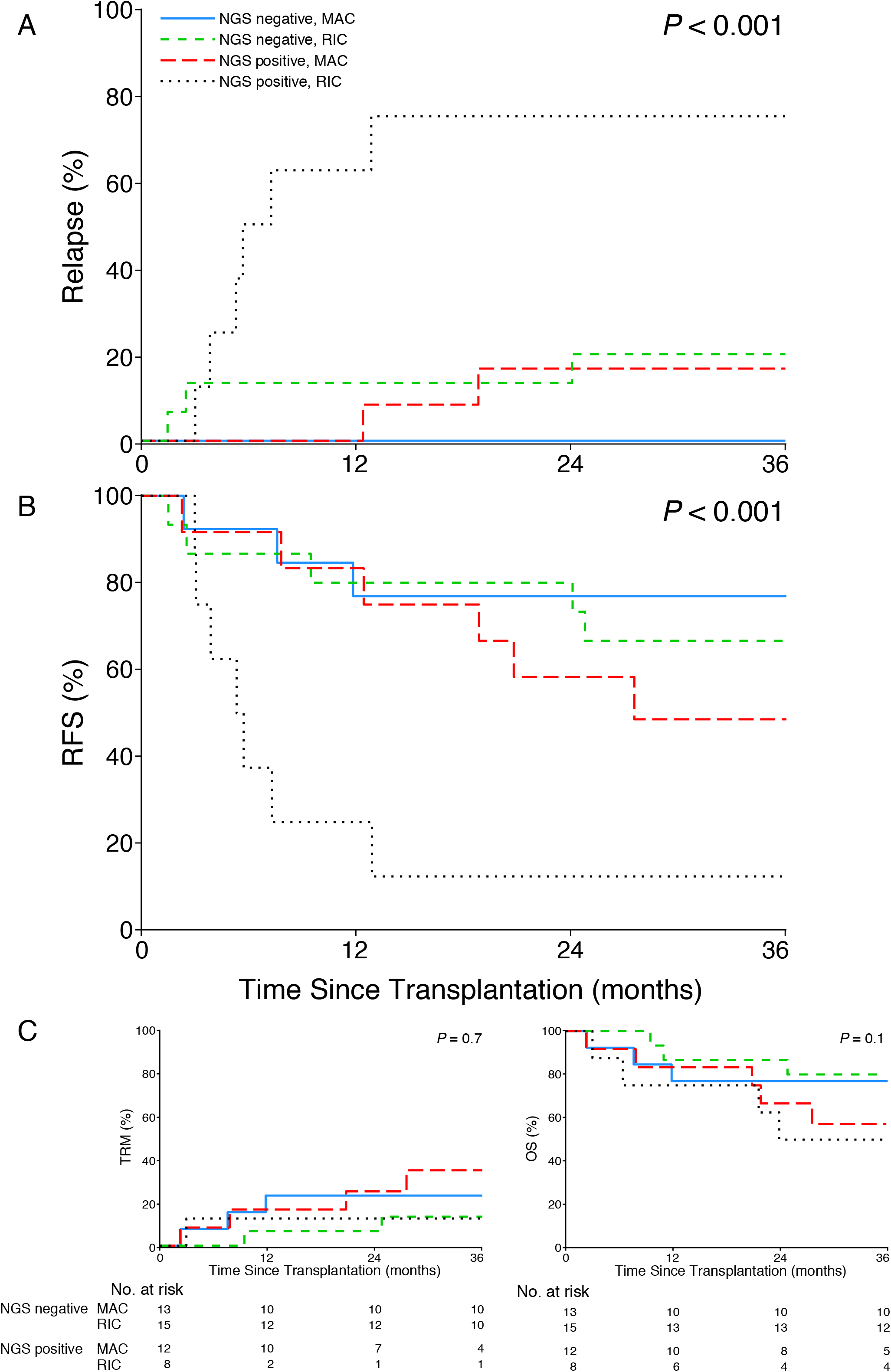
Impact of conditioning intensity and 10-gene mutational status on MDS patient clinical outcomes. (A) Differences in rates of relapse were identified between subgroups defined by conditioning intensity (reduced intensity conditioning [RIC] or myeloablative [MAC]) and mutational status (*P* < 0.001), with the highest rate occurring in 10-gene next-generation sequencing (NGS) positive MDS patients receiving RIC. (B) In patients with no mutations detected within the 10-gene region (NGS negative), no difference in relapse-free survival (RFS) was observed between conditioning intensities (3-year RFS, 67% RIC vs 77% MAC, *P* = 0.63). However, in NGS positive patients, RFS was significantly worse in those who received RIC (3-year RFS, 13% RIC vs 49% MAC, *P* < 0.001). (C) No difference in transplant-related mortality (*P* = 0.7) or overall survival (*P* = 0.1) was observed between subgroups.

For patients testing NGS_g10_ negative, no differences in clinical outcomes between the two conditioning intensity arms were detected, including rates of relapse (3-year relapse, 20% RIC vs 0% MAC, *P* = 0.095), relapse-free survival (3-year RFS, 67% RIC vs 77% MAC, *P* = 0.63), or overall survival (3-year OS, 80% RIC vs 77% MAC, *P* = 0.99) (**Figure 2, Table 2**).

The presence of a NGS_g10_ mutation in the blood prior to conditioning predicted 67% of RIC and 100% of MAC relapses at 24 months, with a specificity of 100% and 64%, respectively (**supplemental Table 4**). Inclusion of patients with poor cytogenetics prior to transplant with NGS_g10_ mutational status improved prediction of relapse at 24 months in the RIC group to 89%, with a specificity of 89%, and is associated with significantly increased rates of relapse and decreased relapse-free survival compared to patients receiving MAC (**supplemental Figure4**).

### Screening for additional genes does not improve test performance

The mutational spectrum of MDS is complex, with many additional genes recurrently mutated beyond the 10 AML-associated genes described above.^7,11,12^ Therefore, we expanded our testing to also cover regions in an additional 19 genes including those commonly found in MDS at diagnosis *(ASXL1, BCOR, CBL, CUX1, DNMT3A, ETV6, EZH2, GATA2, KRAS, PHF6, PPM1D, PTPN11, SETBP1, SRSF2, STAG2, TET2, U2AF1, WT1, ZRSR2)*. This resulted in the detection of an additional 82 mutations (136 in total), with a median VAF of 0.9% and a median of 2.5 variants (range 1-12) per patient (**supplemental Figure 2** and **supplemental Table 3**). Expanding testing (total of 29 genes) resulted in the majority of patients having a mutation detectable prior to conditioning (80% MAC and 78% RIC).

We examined if testing for mutations in a larger number of genes would help better risk stratify patients classified as NGS_g10_ negative (**Figure 3, supplemental Figure 5**). The inclusion of *DTA (DNMT3A, TET2*, and *ASXL1)* genes, known to be associated with age-related clonal hematopoiesis^13,14^, resulted in reclassification of 21% (n=6) of NGS_g10_ negative patients to positive. Similar to what we observed in AML patients^10^, samples testing positive for only *DTA* mutations showed no difference in relapse rates or overall survival compared with those testing negative. Furthermore, while inclusion of *DTA* variants marginally improved the sensitivity of NGS mutational status for predicting relapse at 24 months (from 73% to 82%), it had a detrimental impact on specificity (78% to 35%, **supplemental Table 4**).

**Figure 3.**
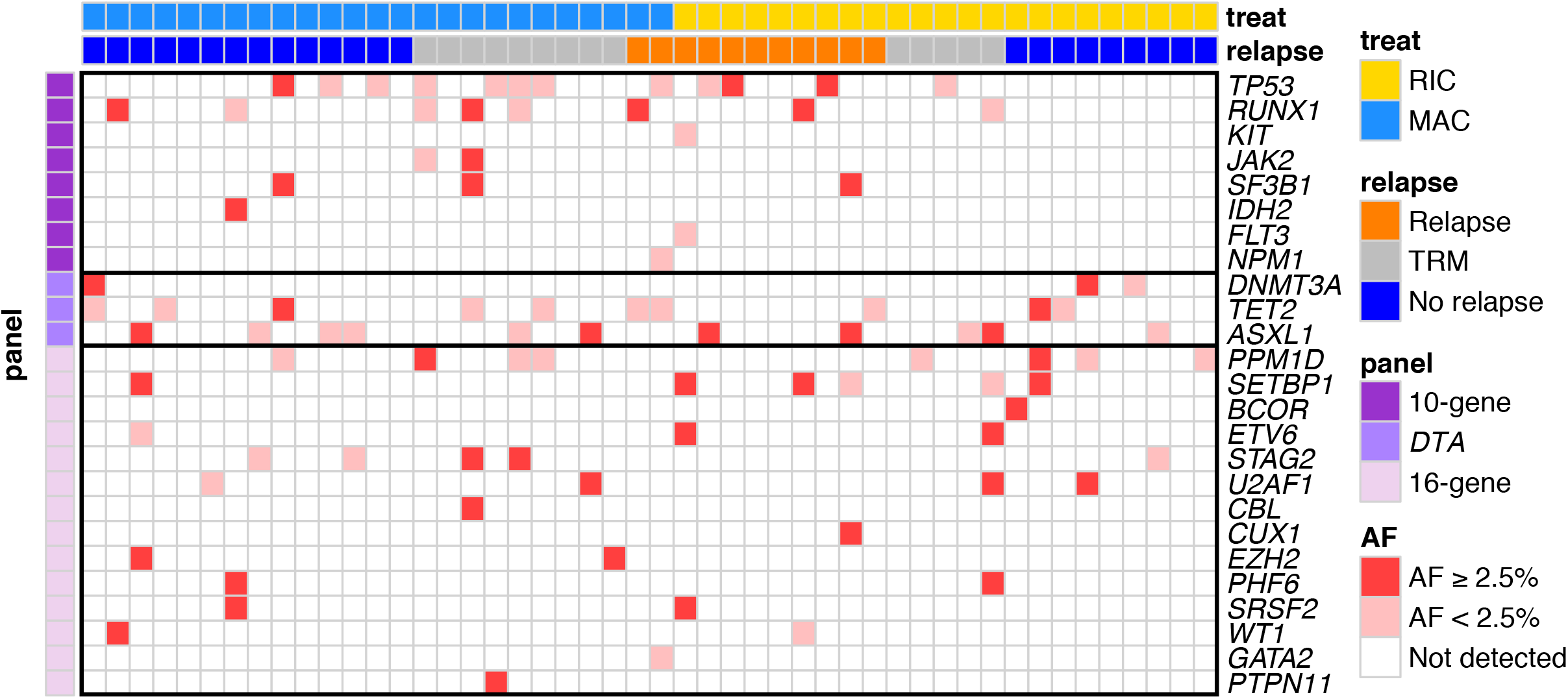
Mutational spectrum of MDS patients prior to conditioning. The heatmap displays mutations detected in the preconditioning blood of MDS patients across 29 gene regions. Patients are displayed in columns and grouped by conditioning intensity (reduced intensity conditioning [RIC] or myeloablative conditioning [MAC]) and clinical outcome (relapse, no relapse, or treatment related mortality [TRM]). Genes are displayed in rows and sorted by diagnostic panel groupings, including 10-gene (prognostic in Acute Myeloid Leukemia), *DTA* (associated with age-related clonal hematopoiesis), and 16-gene (commonly mutated in Myelodysplastic Syndrome). The presence of a mutation within a gene is denoted in the heatmap, with the color corresponding to the highest allele frequency (AF) within each gene per patient.

Inclusion of 16 MDS-associated genes in the testing strategy resulted in reclassification of 43% (n=12) of NGS_g10_ negative patients to positive, but also provided no additional prognostic significance (**Figure 3, supplemental Figure 5**). No relapses were observed in patients testing positive only for these 16-genes regardless of conditioning intensity and only one death was observed (TRM in the MAC group). Additionally, while inclusion of these additional gene regions provided no additional sensitivity for predicting relapse at 24 months, as with *DTA* variants it also reduced specificity (**supplemental Table 4**).

### Variant allele frequency does not provide prognostic significance

Since expanding testing to include all 29 gene regions resulted in the majority of patients having a mutation detected, we examined the impact of defining as positive only those patients with at least one mutation detectable at a higher level (variant allele frequency, VAF, ≥ 2.5%). This resulted in reclassification of 7 NGS_g10_ negative and 5 NGS_g10_ positive patients, resulting in 46% of patients (n=22) being high VAF NGS positive. However, unlike NGS_g10_ positive patients, no significant difference was observed for rates of relapse, relapse-free survival, or overall survival in high VAF NGS positive versus NGS negative patients (**supplemental Figure 6A**). Further stratification of patients by conditioning intensity revealed that 48% of MAC (n=12) and 43% of RIC (n=10) patients were high VAF NGS positive. Higher rates of relapse were observed in high VAF NGS positive patients randomized to RIC versus MAC (3-year relapse, 60% vs 8%; *P* = 0.01), but this effect was driven exclusively by NGS_g10_ mutation status, while no difference was observed in TRM, relapse-free survival, or overall survival (supplemental Figure 6B). Limiting analysis to those testing positive for a NGS_g10_ mutation at or above 2.5% VAF reduced the sensitivity at 24 months from 73% to 45% (**supplemental Table 4**).

### MDS patients with excess blasts

Although the requirement for inclusion in the BMT-CTN 0901 study was less than 5% marrow myeloblasts and no leukemic myeloblasts in blood on morphological analysis at the time of pre-transplant assessment, a total of 22 of the 54 MDS patients (41%) treated on the BMT-CTN 0901 trial had an initial diagnosis of refractory-anemia with excess blasts (RAEB) I or II (ie: between 5-19% blasts). The median time from diagnosis to alloHCT in these patients was 6 months (range 3 – 129 months). These patients accounted for 19 (40%) of the available blood samples from prior to conditioning (9 MAC, 10 RIC). Survival for the 19 RAEB patients with samples available was 74%. Seven of these samples tested positive for NGS_10g_, for which survival was 0% for RIC (n=2, relapse) and 60% for MAC (with 1 death from each relapse and TRM) treated patients. NGS_10g_ positive patients accounted for 4 of 6 relapses observed in the RAEB cohort, the other two patients both had high-risk cytogenetics with a monosomal karyotype that would not be detectable using this NGS assay.

## Discussion

Here we present the impact of genetic mutations detected prior to randomization to RIC or MAC, identified without knowledge of the mutations originally present at diagnosis, on transplant outcomes in MDS patients. Detection of any mutation within regions of 10 genes, previously shown to have utility at the same timepoint in AML patients^10^, was associated with increased rates of relapse and decreased relapse-free survival and overall survival. Many of these genes (including *TP53, RUNX1, JAK2*, and RAS pathway genes) have shown prognostic significance in other MDS studies^7-9^ or are indicative of potential leukemic progression *(FLT3, NPM1)*. The prognostic significance of these mutations on relapse was dependent on conditioning intensity, with higher rates of relapse and lower relapse-free survival observed in patients randomly assigned to receive RIC compared to MAC.

This study provides evidence that the benefit of MAC versus RIC in reducing relapse is found in MDS patients with detectable mutations in a set of 10 high-risk genes prior to alloHCT. In the 58% of MDS patients testing negative, no difference between conditioning arms was seen for relapse, relapse-free survival, or overall survival. It was already well established that detection of mutations in *TP53* prior to alloHCT in MDS patients was associated with increased relapse and decreased survival^7,9^, and that myeloablative conditioning was unable to overcome these risks^9^. In our series, 12 of 48 subjects had a *TP53* mutation, although 9 (75%) of these were below the cut-off (VAF <2.5%) used in previous studies (mean VAF 1.3%, range 0.1% to 19.74%). Four of these subjects relapsed (1 of 8 in MAC group, 3 of 4 in RIC group), five died of transplant-related complications (4 of 8 in MAC group, 1 of 4 in RIC group) and three survived without relapse (all in the MAC group). All three *TP53* mutated survivors who did not relapse had a pre-alloHCT VAF of less than 5% (**Figure 3**). While the size of this cohort limits extrapolation, it is possible that the prognostic implications of *TP53* mutation detection in MDS patients prior to alloHCT and the impact of conditioning intensity differs from that reported previously when those variants are found at a level below the limit of detection of routinely used NGS assays. In addition to VAF, other factors such as persistence since initial diagnosis, development during therapy, mutation type, functional classification, and zygosity are likely to be important in determining the relapse risk associated with detecting a *TP53* mutation.^15-22^

While detection of high-risk genomic features prior to alloHCT was strongly associated with increased relapse and decreased relapse-free survival and these outcomes could be improved with myeloablative conditioning, unlike in AML patients^10^ we did not detect an overall survival advantage for conditioning intensification in those testing positive. This may have been due to limited sample size, predominately non-relapse causes of death in this MDS cohort (67% of deaths, compared with 39% in the previously reported AML cohort), or other factors.

MDS is a genetically heterogenous disease.^1,11,12,23^ As was observed previously^8^, we found that inclusion of a large number of genes resulted in the majority of patients having mutations present prior to transplant. Furthermore, mutations in genes associated with clonal hematopoiesis *(DTA)* or 16 other MDS-associated genes outside of 10-gene mutational status did not improve test performance. Re-analysis of pre-transplant mutational status using only the 10 genes reported here, from another study of MDS patients with <5% myeloblasts prior to alloHCT^8^, confirms our finding of significantly increased relapse rates in patients testing positive and receiving RIC versus MAC. Analysis of larger MDS cohorts using broad NGS panels will have to be performed to define the mutations with greatest prognostic significance at the pre-alloHCT timepoint.

There are several limitations to this work. The modest sample size here offers only a partial sampling of the heterogenous genetics associated with MDS, while differences in pre-transplantation disease burden, biology and prior treatment history further limit exact comparisons. The significance of mutation detection prior to alloHCT may be influenced by history of prior treatment, previous work using the same testing in AML patients included only those treated to cytomorphological remission prior to transplant. Without knowledge of genomics at initial MDS diagnosis we cannot comment on the optimal clinical timepoint or sequencing breadth for testing. Nevertheless, we show here a clear association of pre-transplant genomics with post-transplant relapse and a significant impact of conditioning intensity in reducing that relapse risk.

Unlike in AML, relapse does not account for the clear majority of mortality in patients with MDS undergoing alloHCT.^9^ Two randomized studies of conditioning intensity in MDS have failed to show a difference in survival between MAC and RIC^4,6,24^, although we show here a trend towards improved survival with increased conditioning intensity for those with detectable mutations particularly in the RAEB group (ie: biology and treatment history most analogous to AML). Prior work had shown a benefit for increased conditioning intensity in MDS patients with RAS pathway mutations *(NRAS, KRAS, PTPN11, CBL, NF1, RIT1, FLT3*, and *KIT)* undergoing alloHCT.^9^ As relapse is not the primary cause of death in MDS patients undergoing alloHCT the choice of conditioning intensity is currently dependent on the estimated ability of a patient to tolerate transplant-related toxicity.^25^ AlloHCT remains the only curative therapy for MDS, however given the median age at diagnosis many patients will not be eligible for myeloablative approaches.^26,27^ As it is now possible to determine the genomic basis of disease before and after transplantation, personalized approaches for alloHCT of MDS patients are now conceivable with targeted strategies for those with detectable *TP53*, RAS-pathway (including *FLT3), IDH* and *JAK2* mutations, immune augmentation or other therapies potentially able to supplement the impact of chosen conditioning intensity to minimize relapse risk.^28-30^

In conclusion, we show that in adult MDS patients with less than 5% marrow myeloblasts and no leukemic myeloblasts in blood prior to alloHCT, ultradeep DNA-sequencing for mutations in 10 gene regions previously shown to be high-risk in AML patients could identify a subset of 42% of patients who experienced the majority of post-transplant relapses. In those MDS patients testing positive, myeloablative rather than reduced intensity conditioning could dramatically lower the relapse rate but this benefit was counterbalanced by increased transplant related mortality. This study provides the rationale for clinical trials of personalized post-transplant maintenance for MDS patients based on genetic assessment prior to transplant.

## Data Availability

DNA sequencing FASTQ files are available in the NCBI Small Reads Archive (SRA)

## Acknowledgements

The authors wish to thank R. Coleman Lindsley MD PhD for careful reading of this work.

## Support

This work was supported by the Intramural Research Program of the National Heart, Lung, and Blood Institute (NHLBI) of the National Institutes of Health (NIH) and by grants U10HL069294 and U24HL138660 to the Blood and Marrow Transplant Clinical Trials Network (BMT CTN) from the NHLBI and the National Cancer Institute (NCI). The CIBMTR registry is supported primarily by the U24-CA76518 from NHLBI, NCI and the National Institute of Allergy and Infectious Diseases and from HHSH234200637015C (HRSA/DHHS) to the Center for International Blood and Marrow Transplant Research. This study utilized BMT CTN 0901 research materials, biospecimens and clinical trial data, provided by the BMT CTN. The content is solely the responsibility of the authors and does not necessarily represent the official views of the NIH.

This work utilized the NHLBI Sequencing and Genomics Core and the computational resources of the NIH HPC Biowulf cluster (http://hpc.nih.gov).

## Author Contributions

**Concept and design:**

Dillon, Gui, Hourigan

**Acquisition, analysis, or interpretation of data:** Dillon, Gui, Ghannam, Li, Licon, Alyea, Bashley, Devine, Fernandez, Giralt, Hamadani, Howard, Maziarz, Porter, Warlick, Pasquini, Scott, Horwitz, Deeg, Hourigan

**Drafting of the manuscript:** Dillon, Gui, Hourigan,

**Critical revision of the manuscript for important intellectual content:** Dillon, Gui, Logan, Fei, Ghannam, Li, Licon, Alyea, Bashley, Devine, Fernandez, Giralt, Hamadani, Howard, Maziarz, Porter, Warlick, Pasquini, Scott, Horwitz, Deeg, Hourigan

**Statistical analysis:** Dillon, Gui, Logan, Fei, Licon

**Supervision:** Hourigan

